# Publishing at any cost: a cross-sectional study of the amount that medical researchers spend on open-access publishing each year

**DOI:** 10.1101/2020.12.16.20247643

**Authors:** Mallory K. Ellingson, Xiaoting Shi, Joshua J. Skydel, Kate Nyhan, Richard Lehman, Joseph S. Ross, Joshua D. Wallach

**Author notes:** **Corresponding author:** Joshua D Wallach, MS, PhD, Assistant Professor, Department of Environmental Health Sciences, Yale School of Public Health, 60 College Street, 4^th^ Floor, Room 411, New Haven, CT, 06510 USA, **Twitter:** @JoshuaDWallach.

## Abstract

**Objective:** To estimate the financial costs paid by individual medical researchers from meeting the article processing charges (APCs) levied by open access journals in 2019. To investigate the emotional burden to researchers using a novel metric (the APC Twitter Whinge Score).

**Design:** Cross-sectional analysis.

**Data sources:** Scopus was used to generate two random samples of researchers, the first with a senior author article indexed in the ‘Medicine’ subject area (i.e., general researchers) and the second with an article published in the ten highest impact factor general clinical medicine journals (i.e., high-impact researchers) in 2019. For each researcher, Scopus was used to identify all first and senior author original research and review articles published in 2019. Researcher and journal information was obtained from Scopus, institutional profiles, Journal Citation Reports, publisher databases on APCs, the Directory of Open Access Journals, and individual journal websites. Twitter searches were conducted to identify and classify APC-related tweets using a novel APC Twitter Whinge Score.

**Main outcome measures:** Median APCs paid by general and high-impact researchers for all first and senior author research and review articles published in 2019; additionally, we examined median APCs paid by researcher gender, affiliation, training, and geographic region. APC Twitter Whinge Score.

**Results:** There were 241 general and 246 high-impact researchers identified as eligible for our study. In 2019, the general and high-impact researchers published a total of 914 (median 2, interquartile range 1-5) and 1471 (4, 2-8) first or senior author research or review articles, respectively. 42% (384/914) of the articles from the general researchers and 29% (428/1471) of the articles from the high-impact medical researchers were published in fully open access journals. The median total APCs paid by general researchers in 2019 was $191 [£150] (interquartile range $0-$2500 [£0-£1960]) and the median total paid by high-impact researchers was $2900 [£2274] (interquartile range $0-$5465 [£0-£4285]); the maximum paid by a single researcher in total APCs was $30115 [£23610] and $34676 [£27186], respectively. There were no differences in total APCs paid by gender, affiliation, or training. However, high-impact researchers from the Region of the Americas had a lower median total APCs paid than those from other regions ($1695, interquartile range $0 - $3935) [£1329, £0-£3085] vs. $4800, $1888-$8290) [£3763, £1480-£6500]; P<0.001). Among a sample of 195 APC-related tweets in 2019, 121 (62.1%) were publicly resentful (with or without sweary language) of APCs, scoring in the highest two categories of the APC-related Twitter Whinge Score.

**Conclusions:** Medical researchers in 2019 were found to have paid between 0 [£0] and $34676 [£27186] in total APCs. As journals with APCs become more common, it is important to understand the cost to researchers, especially those who may not have the funding or institutional resources to cover these costs, or we risk creating a pay-to-publish system that favors well-resourced authors from well-resources institutions and areas of the world. We also present evidence that these APCs may cause emotional damage to researchers, causing them to divert effort to moaning on Twitter. We postulate that behind this behavior may lie hidden harmful cycles of personal penury, domestic argument, insomnia, poor work relationships, inadequately prepared coffee, and even the possible use of alcohol before tweeting.

**What is already known on this topic:**

- Over the past 20 years, a new model of scientific publishing has emerged that relies on digital publication rather than print distribution – open access publishing.
- Open access publishing has shifted part of the financial costs of publishing from academic institutions to individual researchers and their funders, who are responsible for article processing charges that can average $2000 (£1568) to $3000 (£2352) per article.
- In additional to the potential financial costs of publishing, anecdotal evidence suggests that article processing charges may lead to emotional damage to researchers, causing them to divert effort to moaning on Twitter.

**What this study adds:**

- Medical researchers could be paying as much as $34676 [£27186] in total article processing charges for their first and senior research and review articles each year.
- The majority of article processing charge related tweets are publicly resentful (with or without sweary language) of these journal fees, scoring in the highest two categories of the Twitter Whinge Score.
- As journals with article processing charges become more common, it is important to understand the burden – financial and emotional - on researchers, especially those who may not have the funding or institutional resources to cover these costs.

## INTRODUCTION

Publications in peer-reviewed journals are currency in the academic world, and are often viewed as a proxy for productivity, competency, and prestige. With over 15 million publishing scientists across the world,^1,2^ the pressure to publish has only risen, as has the importance of publications for employment, promotion, and tenure.^3^ Over the past decade, there has been a striking growth in the number of scientific articles published per year, with nearly 2.5 million scientific articles published in 2018 alone.^4^ The sheer quantity of scientific research being published, the shift to predominantly electronic publishing, and a broad movement to make scientific research more transparent has wrought a dramatic change in the landscape of scientific publishing.^5,6^

Currently, the primary mechanism for the publication of scientific articles is through peer-reviewed journals. These journals have operated using a subscription model, generally owned and managed by a professional society or a medical publisher. Under this model, the cost to individual researchers, either to access articles or publish their own research in the journals, is minimal (although they contribute substantial in-kind effort through peer and editorial review). Instead, institutions pay subscription fees, which can reach millions of dollars for larger publishers, to gain access to articles for the institution’s affiliates.^7,8^ Receipt of the *BMJ* is a compulsory benefit imposed on members of the British Medical Association in return for an annual membership fee.

However, over the past 20 years, a new model of scientific publishing emerged that relied on digital “publication” rather than print distribution – open access publishing.^5^ Generally, open access journals forgo subscriptions for their online content and instead make research available to scholars without institutional subscriptions and to the general public. With no revenue from subscriptions (or from prescription drug advertisements), some open access journals established a new business model built around article processing charges (APCs).^5^ Compared to the subscription journals, the APC model has shifted part of the financial burden of publishing from academic institutions to individual researchers and their funders, who are responsible for APCs that average $2000 (£1568) to $3000 (£2352) per article.^9,10^ In this way, everyone can eat the picnic, but those who provide the food do so at their own cost and are additionally charged for bringing it to the picnic site.

With almost 5000 open access journals following the APC business model,^11^ researchers are increasingly having to consider if and how they can afford to publish their research in open access journals with the limited pool of funds available. While the vast majority of medical researchers are supportive of the concept of open access publishing, over half listed financial barriers as the primary reason they would choose not to publish in open access journals.^12^ Although APCs can be covered with funds from research grants or by funders directly, not all research is grant funded, the structure and amount of funding that comes from grants can vary by field, and the ability or willingness of funders to cover APCs differs by region.^10,13^ Additionally, early career researchers or under-represented minority (URM) researchers may have more limited access to grant funding or institutional funds to cover APCs, as do researchers in less lucrative clinical fields like primary care and public health.^14–16^ While fee waivers are sometimes granted to researchers in low- and middle-income countries or without funding, discounted APCs may still be prohibitive for many researchers.^17,18^

If financial barriers play such a substantial role in scientists’ decisions on where to publish, it is important to investigate the potential financial costs of publishing on individual medical researchers. Therefore, we aimed to estimate how much individual medical researchers spend on APCs over the course of a year for both a general sample of medical researchers as well a sample of researchers who published in the highest impact factor clinical medicine journals in 2019. To measure the emotional burden of APCs on researchers, we also evaluated APC-related tweets using a novel metric (The APC Twitter Whinge Score).

## METHODS

### Part 1: Estimating article processing charges

#### Study design and sample

We conducted a cross-sectional analysis of two random samples of medical researchers to obtain estimates of the average amount of money that individual researchers spend on APCs each year. We used Scopus to identify 250 general medical researchers and 250 medical researchers who published at least one article in one of the ten highest impact factor general clinical medicine journals.

#### Generating a sample of general and high-impact researchers

First, we downloaded the first 20000 English language research or review articles published in a journal indexed in the Scopus subject area of ‘Medicine’ in 2019 (**Figure 1)**, the maximum data export permitted through the Scopus portal. Next, we used Scopus to identify all research or review articles published in the top ten highest impact factor clinical medicine journals in 2019 (according to the Journal Citation Report (JCR):^19^ *New England Journal of Medicine (NEJM), Lancet, Journal of the American Medical Association (JAMA), British Medical Journal (BMJ), JAMA Internal Medicine, Annals of Internal Medicine, PLOS Medicine, BMC Medicine, Mayo Clinic Proceedings*, and *Canadian Medical Association Journal*). For each sample, we used a random number generator to select 500 articles. However, given the broad nature of the search utilized, some articles randomly selected did not fall under the category of “general clinical medicine,” or were other article types misclassified as research or review articles. Therefore, the first 250 articles determined to be eligible were retained for each sample (i.e., 250 general medicine articles and 250 high-impact medicine articles).

**Figure 1.**
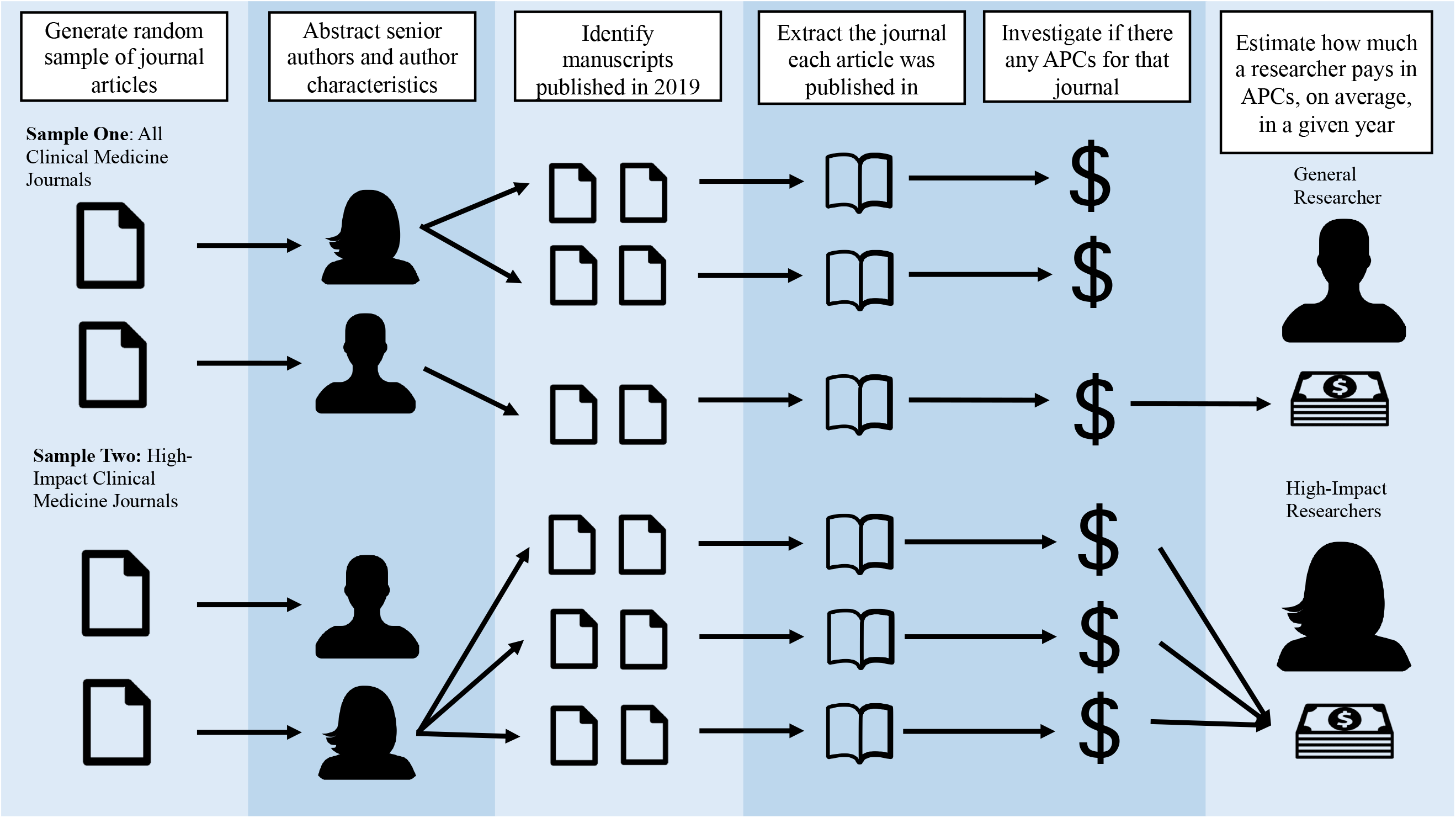
A visualization of the sampling and data abstraction approach

Next, we identified the senior-most (i.e., last) author of each research or review article contained in the sample (hereafter, index researcher). Potential duplicate researchers were verified and removed through a Scopus and/or Google scholar search of the researcher’s name. If authorship was listed as a group, without any designated individuals, the manuscript was excluded from the sample. If a group authorship was listed as the senior author, the senior-most individual author on the article was used.

#### Data collection

Three investigators (MKE, XS, JJS) independently abstracted data, and to ensure data quality, approximately 20% of each sample was abstracted in duplicate to verify consistency. All uncertainties were discussed with a fourth investigator (JDW). All data abstraction and validation were conducted between April 22^nd^ and July 22^nd^, 2020.

#### Researcher Information

For the index researchers in both samples, we used the researcher’s Scopus profile to collect the researcher’s name, affiliation, geographic region (based on the six World Health Organization [WHO] regions),^20^ H-index, year of first publication, and research field. Research field was collected from the subject area tags listed on the researcher Scopus profile. For each index researcher, we conducted a Google search of the researcher’s name and screened the first 10 pages to identify an institutional researcher profile. If a researcher profile was available, we also abstracted researcher gender, if clearly indicated in the profile or through identification of gender pronouns, and training (a doctor of medicine (MD), with or without other degrees; a doctor of philosophy (PhD), with or without other degrees (excluding MD); or any other degrees).

#### Identification of first and senior author publications in 2019

Using the Scopus profile of each index researcher, we identified all of the articles published in 2019 where the index researcher was listed as either the first or senior author. Articles on which the index researcher was a middle author (no matter if 3 authors were listed or 20) were excluded, as we assumed the index researcher would not have paid any associated APC as a middle author. For each article, we abstracted the corresponding journal’s title and determined whether the article was marked as open access in Scopus.^21^

#### Journal characteristics and article processing charges

We used JCR to determine the 2018 journal impact factor for each unique journal. Next, we identified the journal publishing model (open access, hybrid, or subscription-based) and the APCs for each journal. A hybrid journal was defined as a traditional subscription-based journal with a fee-based open access publication option.^22^ To ascertain APCs, we first utilized publisher specific databases,^23–37^ which provide lists of open access and hybrid journals from selected publishers and their associated APCs. If a journal could not be identified through a publisher database, we utilized the Directory of Open Access Journals (DOAJ)^11^ to identify if the journal was open access. If listed on DOAJ, the journal was considered open access and the corresponding APC was collected from the provided link to the journal website on DOAJ. If the publishing model of a journal could not be determined from those two sources, we relied on the information provided on individual journal websites. Journals without a clear open access policy (either full open access or a hybrid approach) were considered subscription-based.

We defined the standard APC for an open access journal as the fee associated with publishing in that journal. For hybrid journals, we defined the APC as the fee associated with optional open access publication. We did not include additional fees not associated with open access publication, such as charges for color printing or reprints, as part of the APCs. If an APC for a given journal was based on word count or page limits, we approximated the standard APC using an average article (3500 words) or page count (8 pages).^38^ In addition to the standard APC, we collected the minimum APC for any journal with multiple APC options. The minimum APC was defined as the lowest APC a researcher could pay given any discounts publicly listed by the journal on the journal website or in the publishing database or different licensing options (e.g. institutional or author membership discounts or commercial versus non-commercial licenses).

#### Statistical Analysis

Using descriptive statistics, we characterized the sample of both the general researchers and high-impact researchers, including gender, affiliation, training, geographic region, and seniority (based on H-Index and length of the researcher’s career). Length of the researcher’s career was approximated by subtracting the year of the index researcher’s first publication from 2020.

Next, we calculated the median (interquartile range) APCs paid by index researchers in 2019 for both groups. To do this, for each index researcher, we calculated the maximum total APCs paid in 2019 by assuming that an APC was paid by the index researcher if they were the first or senior author of an article. If an index researcher’s article was published in an open access journal with an APC listed, we assumed that the APC was paid without any discounts or waivers. For any of the index researcher’s articles published in either hybrid or subscription-based journals, we assumed no APC was paid. Lastly, we also calculated the proportion of articles published in open access journals.

We used the Mann Whitney U test or Mood’s test as appropriate to compare median APCs paid per index researcher by the characteristics noted above. For comparisons of APCs paid by H-Index and length of the researcher’s career, we compared researchers above and below the median H-Index and across quartiles in each sample, respectively. Any unknown values were considered as missing. US dollar amounts were converted to British pound sterling using the average exchange rate for 2019.^39^ All data analyses were conducted in R Version 3.6.1 and used a threshold for statistical significance of 0.05.

#### Sensitivity analyses

We repeated the analyses above assuming that index researchers paid: (1) the minimum publicly listed APCs, (2) the APCs for articles published in hybrid journals and classified as “Open Access” on Scopus, (3) APCs for only their first author articles, and (4) APCs for only their senior author articles.

### Part 2: The APC Twitter Whinge Score

To measure the emotional burden that APCs can have on researchers, we also developed and tested a novel metric - the APC Twitter Whinge Score (**Box 1**). This scoring system uses a highly scientific process to manually screen and evaluate the language used to describe APCs on Twitter. Nine advanced searches were conducted on Twitter to identify Tweets posted between January 1, 2019 and December 31^st^, 2019, containing the phrases “Article cost”, “Article costs”, “Article fee”, “Article processing costs”, “Article processing fee”, “Article processing charge”, “Publication cost”, “Publication costs”, and “Publication fee”. One author (JDW) screened all search results, identified any APC-related Tweets from individual accounts (i.e., excluding Tweets from organizations or journals/publishers advertising their amazingly low APCs), and characterized the tweets using the APC Twitter Whinge Score.

#### Box 1.

The Article Processing Charge Twitter Whinge Score © ™ ®

| **Box 1. The Article Processing Charge Twitter Whinge Score © ™ ®** |  |
| --- | --- |
| **Publicly grateful** | Tweets praising or defending article processing costs |
| **Calmly resigned** | Tweets that note article processing costs, but neither tip their hand toward positive or negative; factual statements |
| **Publicly resentful** | Tweets containing strong or sarcastic language, expressing concern, anger, or regret towards article processing costs (bonus points: emoji's and GIFs) |
| **Publicly opposed** | Tweets containing swear language, not suitable for publication in a journal |

#### Patient and Public Involvement

This study was an analysis of publicly available, non-clinical data. There was no patient or public involvement in any of the phases of the study, although we expect the public to be broadly supportive of open access publishing since it permits access to information and does not require expensive subscriptions through medical libraries.

## RESULTS

After accounting for duplicate index researchers and non-English publications, our sample included 241 general and 246 high-impact researchers. Among the 241 general researchers, 239 (99.2%) had ‘Medicine’ listed as one of their subject areas on Scopus; all 246 high-impact researchers had ‘Medicine’ listed as one of their subject areas.

### Researcher characteristics

Nearly all of the general researchers were affiliated with academic centers or hospitals (236/241, 97.9%); 62 (25.7%) were based in the Americas, 76 (31.5%) in Europe, and 69 (28.6%) in the Western Pacific region (**Table 1**). An institutional profile could not be identified for approximately one quarter of the researchers (62/241, 26.5%). Among the 179 researchers with an institutional profile, two thirds had an MD (120/179, 67.0%) and 70 (70/179, 39.1%) were women. On average, general researchers had published at least two (median: 2, IQR 0 - 4) first or senior author articles in 2019, had an H-Index of 11 (median: 11.0, IQR 3.0 - 23.0), and had at least a 15-year publication history.

**Table 1.** Characteristics of general and high-impact medical researchers

|  |  |  |
| --- | --- | --- |
|  | **General researchers**<br>**(N = 241)** | **High-Impact**<br>**researchers**<br>**(N = 246)** |
|  | **No. (%)** |  |
| **Gender** |  |  |
| Male | 109 (45.2) | 159 (64.6) |
| Female | 70 (29.1) | 82 (33.3) |
| Unknown/Unavailable<sup>a</sup> | 62 (25.7) | 5 (2.0) |
| **Affiliation** |  |  |
| Academia/Hospital | 236 (97.9) | 213 (86.6) |
| Government | 3 (1.2) | 10 (4.1) |
| Non-governmental or non-profit | 1 (0.4) | 12 (4.9) |
| Industry | 1 (0.4) | 7 (2.9) |
| Other | 0 (0.0) | 2 (0.8) |
| Unknown/Unavailable | 0 (0.0) | 2 (0.8) |
| **Training<sup>b</sup>** |  |  |
| MD | 120 (49.8) | 166 (67.5) |
| PhD only | 58 (24.1) | 61 (24.8) |
| Other degree only | 3 (1.2) | 5 (2.0) |
| Unknown/Unavailable | 60 (24.9) | 14 (5.7) |
| **Region** |  |  |
| African Region | 3 (1.2) | 4 (1.6) |
| Region of the Americas | 62 (25.7) | 144 (58.5) |
| South-East Asia Region | 13 (5.4) | 1 (0.4) |
| European Region | 76 (31.5) | 67 (27.2) |
| Eastern Mediterranean Region | 18 (7.5) | 4 (1.6) |
| Western Pacific Region | 69 (28.6) | 25 (10.2) |
| Unknown/Unavailable | 0 (0.0) | 1 (0.4) |
| **H-Index, median (IQR)** | 11.0 (3.0, 23.0) | 38.5 (22.0, 64.0) |
| **Years since first publication, median (IQR)** | 15.0 (7.0, 25.0) | 22.5 (14.3, 32.0) |
| **Number of articles per author, median (IQR)** |  |  |
| First and senior author articles | 2 (1, 5) | 4 (2, 8) |
| Senior author articles only | 2 (1, 4) | 4 (2, 8) |
| First author articles only | 0 (0, 1) | 0 (0, 1) |
| Corresponding author articles (first or senior) | 1 (0, 2) | 2 (0, 4) |
| MD = doctor of medicine; PhD = doctor of philosophy |  |  |
| <sup>a</sup> Institutional profile was not available or author gender could not be determined through an institutional profile |  |  |
| <sup>b</sup> MD, with or without other degrees; a PhD, with or without other degrees (excluding MD); or any other degrees |  |  |
| <sup>c</sup> Based on Scopus classification |  |  |

The vast majority of high-impact researchers were affiliated with academic centers or hospitals (213/246, 87.6%); 85.8% (211/246) were primarily based in the Americas or Europe (**Table 1**). An institutional profile was identified for almost all (241/246, 98.0%) of the high-impact researchers. The majority of those with an institutional profile (166/241, 68.9%) had an MD and one-third (82/241, 34.0%) of the researchers were women. High-impact researchers had, on average, a publication history of greater than 20 years, an H-Index of 38.5 (median: 38.5, IQR 22.0 - 64.0), and had published at least four first or senior author manuscripts in 2019 (median: 4, IQR 2 - 8).

### Article Characteristics

In 2019, the 241 general researchers published 914 first or senior author research or review articles in 598 unique journals. The most common journals were *Medicine* (15/914, 1.6%) and the *International Journal of Molecular Sciences* (9/914, 1.0%). Among the 462 journals with a 2018 JCR impact factor, the median impact factor among their articles was 2.65 (IQR 1.69-3.86). The 246 high-impact researchers published 1471 original first or senior author research or review articles in 604 unique journals. The most common journals were *New England Journal of Medicine* (60/1471, 4.1%), *The Lancet* (43/1471, 2.9%), and *PLOS Medicine* (36/1471, 2.5%). Among the 537 journals with a 2018 JCR impact factor, the median impact factor among their articles was 5.05 (IQR 3.18-11.05).

Of the 914 first or senior author research or review articles published by the general researchers, 414 (414/914, 45.3%) were indexed in Scopus as open access. There were 384 (41.6%) articles published in an APC-based journal (**Table 2**). Of the 457 (457/914, 50.7%) articles published in journals with a hybrid funding model, 72 (72/457, 15.8%) were indexed as open access. Among the high-impact researchers, 726 (726/1471, 49.4%) of the articles were indexed in Scopus as open access. Just under one-third of all articles were published in an APC-based journal (426/1471, 28.9%). Among the 870 (870/1471, 59.1%) articles published in journals with a hybrid funding model, less than one-third (255/870, 29.3%) were open access.

**Table 2.** Journal funding models for the articles published by general and high-impact researchers

|  |  |  |
| --- | --- | --- |
|  | **No., (%)** |  |
|  | **General researchers** | **High-impact researchers** |
| **Total** | 914 | 1471 |
| Article processing charge-based journals | 384 (42.0) | 426 (29.0) |
| Subscription-based journals | 57 (6.2) | 169 (11.5) |
| Hybrid<sup>a</sup> | 457 (50.0) | 870 (59.1) |
| Unknown | 16 (1.8) | 6 (0.4) |
| <sup>a</sup> A traditional subscription-based journal with a fee-based open access publication option |  |  |

### Article Processing Charges

The journal funding model and any associated APCs could be identified for 94.1% (860/914) of the first or senior research or review articles published by the general researchers and 97.8% (1439/1471) of the articles published by the high-impact researchers. In 2019, the 241 general and 246 high-impact researchers paid an estimated total of $497716 (£390209) and $1067869 (£837209) in APCs, respectively, for their first and senior author articles. Although the median APCs paid by general clinical medical researchers was $191 (IQR $0 - $2500) [£150, £0 - £1960], one researcher was estimated as having paid $30115 (£23610) in APCs (**Table 3, Figure 2)**. The median total APCs per researcher in the high-impact sample was $2900 (IQR $0 - $5465) [£2274, £0 - £4285]; one researcher was estimated as having paid as much as $34676 (£27186) in APCs.

**Table 3.** Article processing charges for all first and/or senior research and review articles published in 2019

|  |  |  |
| --- | --- | --- |
|  | **Median (Interquartile range)** |  |
|  | **General researchers (n = 241)** | **High-impact researchers (n = 246)** |
| **Standard APCs paid per year, \$** | 191 (0, 2500) | 2900 (0, 5465) |
| First author articles only | 0 (0, 0) | 0 (0, 0) |
| Senior author articles only | 0 (0, 0) | 2800 (0, 5181) |
| **APCs paid per year (including hybrid journals)<sup>a</sup>, \$** | 739 (0, 3950) | 5000 (0, 10879) |
| **APCs paid per year (minimum)<sup>b</sup>, \$** | 0 (0, 2500) | 2600 (0, 5465) |
| APCs = article processing charges |  |  |
| <sup>a</sup> A traditional subscription-based journal with a fee-based open access publication option |  |  |
| <sup>b</sup> The minimum APCs paid is defined as the lowest possible APC an author could have paid given the discounts, membership options or licensing options listed on a journal website. |  |  |

**Figure 2.**
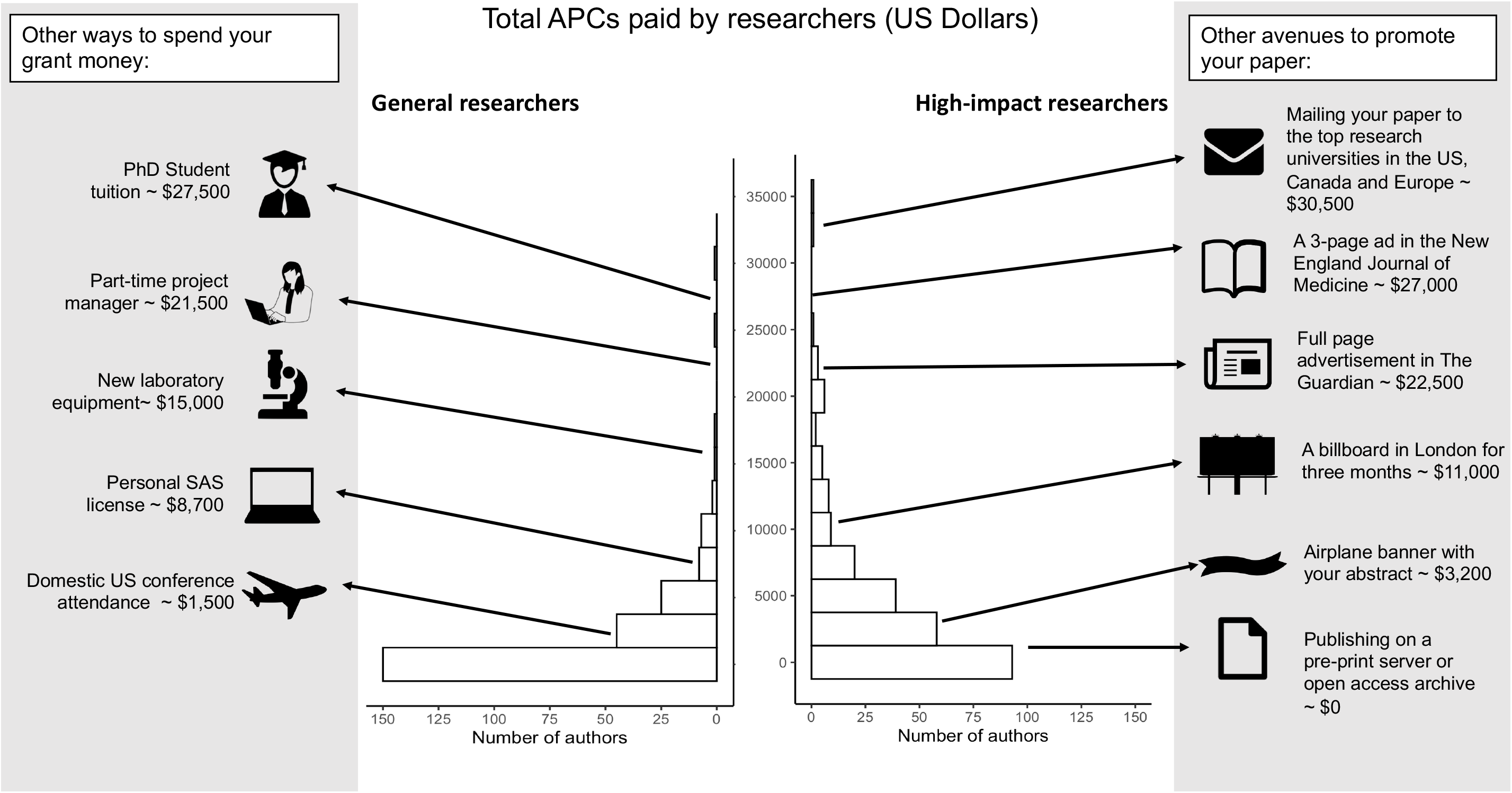
Distribution of research spending on article processing charges in 2019

In sensitivity analyses, after including potential discounts on standard APCs, the minimum listed APCs general researchers could have paid for their first and senior author publications in 2019 was $0 (IQR: $0 - $2500) [£0, £0 - £1960] (**Table 3**). However, researchers in the high-impact sample would have paid approximately $300 less on average (median: $2600, IQR $0 - $5465) [£2038, £0 - £4285]. If all researchers paid the APCs for their first and senior open access published in hybrid journals (as opposed to the articles being made available through delayed open access due to funder requirements, at the discretion of the journal, or through other mechanisms such as self-archiving) the median total APCs paid by the general and high-impact researchers would have been $739 (IQR $0 - $3950) [£579, £0-£3097] and $5000 (IQR $0 - $10879) [£3920, £0-£8529], respectively.

The estimated median total APCs paid did not vary across index researcher gender, training, H-index, and years since first publication (**Table 4)**. However, high-impact researchers in the Region of the Americas did have lower median total APCs per researcher than those in other regions of the world (Region of the Americas: $1695, IQR $0-$3935 [£1329, £0 - £3085] vs. Other regions: $4800, IQR $1888-$8290 [£3763, £1480 - £6500]; p <0.001).

**Table 4.**
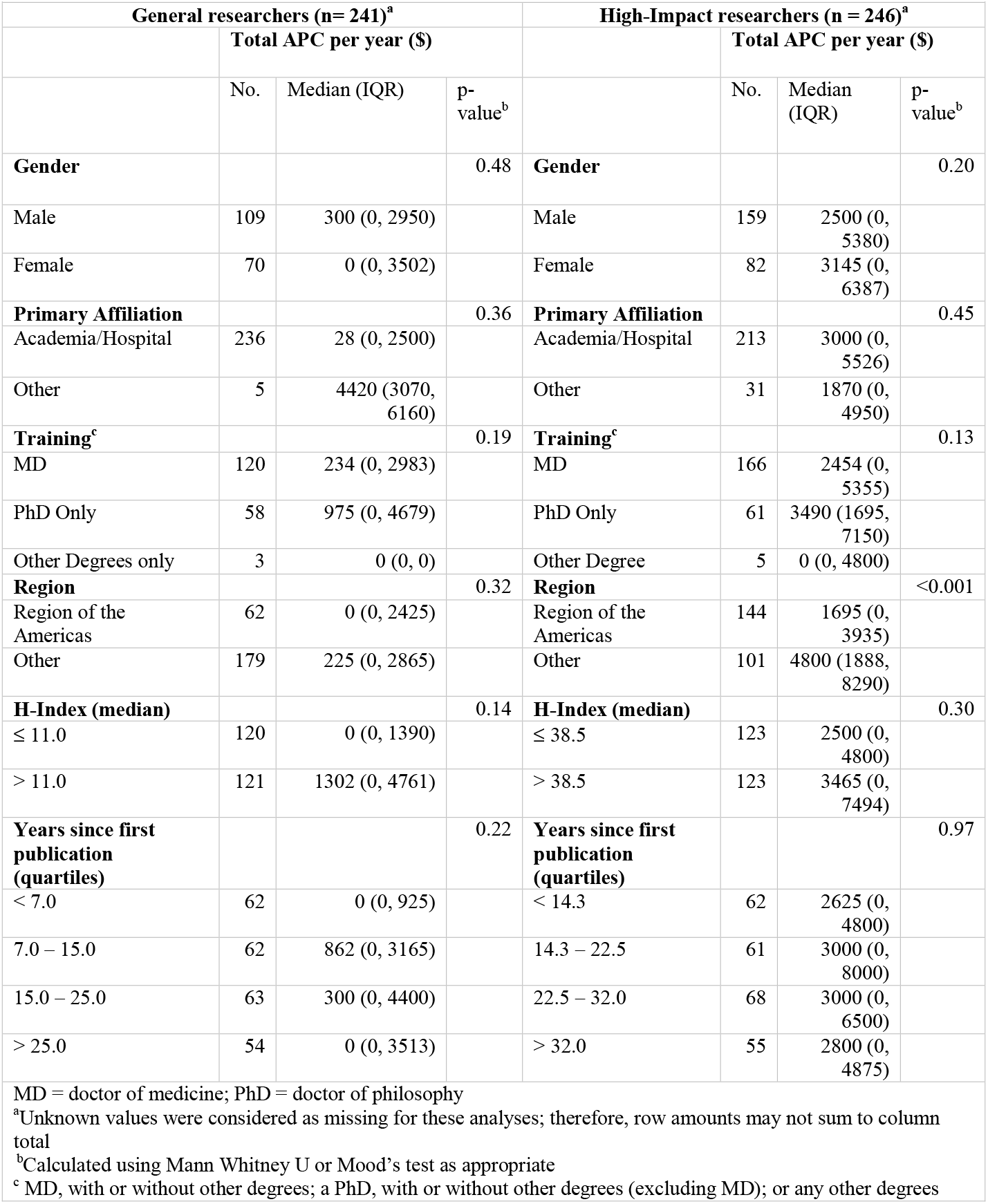
Standard article processing charges for first and senior research and review articles, across researcher characteristics

### The APC Twitter Whinge Score

Among the sample of 195 APC-related tweets posted by individuals in 2019, the majority (118/195, 60.5%) were classified as publicly resentful of APCs (**Figure 3**). There were two (2/203, 1.0%) additional tweets that described APCs using sweary language (although, we can only assume that the authors meant “Why The Fee!?” when they wrote WTF).

**Figure 3.**
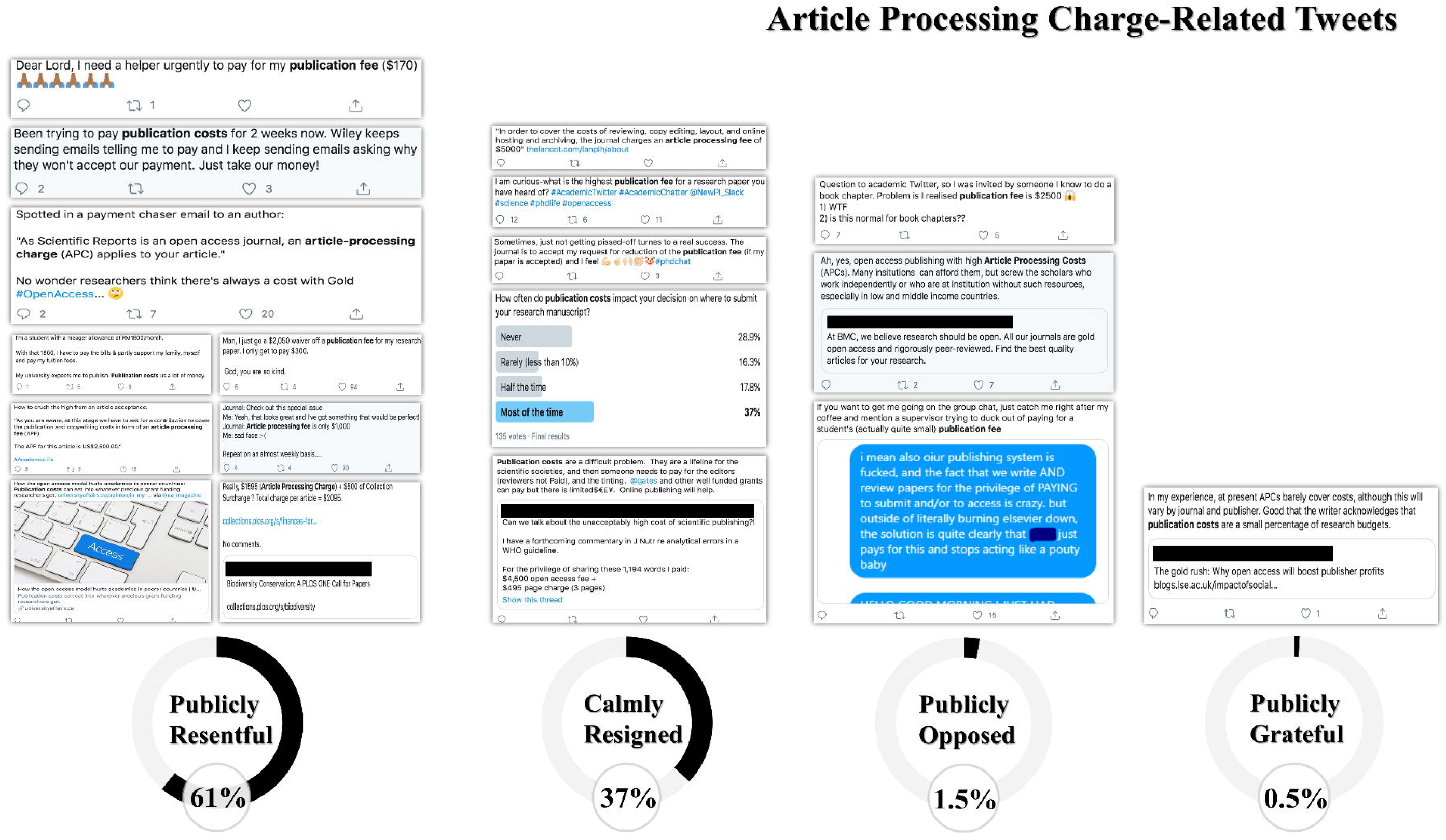
The Article Processing Charge-Related Twitter Whinge Score

## DISCUSSION

In this cross-sectional study, 241 and 246 randomly selected general and high-impact medical researchers published a median of 2 and 4 first or senior author research or review articles in 2019, respectively. Approximately one-third of the articles across both samples were published in journals that required an APC. The median total APCs per general and high-impact researcher in 2019 was $191 [£150] and $2900 [£2274], respectively, with one researcher who may have incurred as much as $34676 in APCs [£27186]. Across both samples, there were no meaningful differences in APCs paid by gender, affiliation, or training. However, in the high-impact sample, researchers from the Region of the Americas had a lower median total APCs paid ($1695) [£1329] than researchers from all other regions ($4800) [£3763]. As open access publishing with APCs becomes increasingly common, it is important to consider the financial (and emotional) implications for individual researchers across different fields, settings, and levels of seniority. Otherwise, we risk creating a pay-to-publish system that favors well-resourced authors from well-resourced institutions and areas of the world. Without a solution that works for all stakeholders, we fear that Twitter will become over-run with tweets whinging about APCs.

Our study suggests that many general and high-impact researchers could have paid thousands of dollars in APCs to publish their first and senior research and review articles in 2019. Across the 487 index researchers in both samples, which represents only a fraction of all biomedical researchers actively publishing in 2019, the total estimated APCs was approximately $1500000 [£1176000]. Given that general researchers published a median of 2 first or senior articles per year (potentially in lower impact factor journals with smaller APCs)^40,41^ it may not be surprising that the median total APCs per researcher was relatively low ($191) [£150]. However, among high-impact researchers, who published a median of 4 first or senior research or review articles in 2019, the median total APCs per researcher was $2900 [£2274]. This suggests that these researchers paid an APC for one of every four of their first or senior author articles. Moreover, if we extrapolate our findings, individual researchers could spend a total of $116000 [£90944] on publication costs over a 40-year career.

The rise of the APC-centered open access publishing model poses a number of serious challenges for researchers.^18,42^ Approximately one-third of the first or senior research and review articles published by the general and high-impact researchers were published in an open access journal that required an APC. Although not all open access journals charge APCs, approximately 50% of all articles that are published open access are published in journals that do.^43^ When grant money or institutional discretionary funds are used to cover APCs, as is the case for approximately 80% researchers in the health, biological, and life sciences,^10^ fewer resources are available for other research-related expenses.^42^ For instance, the $2900 [£2274] median amount spent by researchers in our high-impact sample could support the attendance of multiple individuals at a conference or a critical piece of research equipment (or a number of other radical publication options outlined in **Figure 2**). Moreover, for researchers spending tens of thousands of dollars a year on APCs, these funds could have covered the tuition of a graduate student or the partial salary of a postdoctoral fellow. Second, the amount of APCs has risen dramatically in recent years – at a rate nearly three times that of the expected inflation rate.^13,44^ These increases have raised questions about whether APCs actually reflect the cost of publishing or if publishers are driven by primarily financial motives.^9,42^ While there does not appear to be a quality difference between subscription-based and open access journals,^5,45^ there is some evidence that journals with higher APCs are perceived to be higher impact.^41,45^

Lastly, the amount of APCs can be prohibitive to many researchers, either based on field^10^, seniority^14^, disparities in research funding^15,16^, or setting.^18,42,46^ For instance, evidence suggests that researchers from countries with gross domestic products (GDP) lower than $25000 [£19600] are more likely to pay APCs out of personal funds compared to researchers from countries with GDPs higher than $25000 [£19600].^10^ It is important to note that certain journals grant fee waivers to researchers from low- and middle-income countries or to researchers without funding to support publication. However, many researchers may be unaware of the specific journals that do provide waivers.^17^ Furthermore, journal waivers do not necessarily address all of the inequities imposed by APCs. For early career researchers (i.e. the first, second, third, and senior authors on this manuscript!), with no established grant funding or accumulated discretionary funds (i.e., the first, second, third, and senior authors on this manuscript!), even discounted APCs can be prohibitive.

As open access publishing becomes the norm, numerous opportunities exist to address the disadvantages that may prevent many researchers from paying for APCs. At the journal level, increased transparency may be necessary to inform researchers from low- and middle-income countries or at early stages of their careers about the waivers that are available. It is also critical that funders and institutions leverage their influence to restrain the hyperinflation of APCs. In 2018, cOAlition S, an international consortium of research funders, launched ‘Plan S’. This initiative, which aims to make all scientific publications resulting from publicly-funded research immediately available open access^47^, has proposed an APC fee cap.^44,47^ As more scientific research is available open access, institutions can shift resources from subscriptions to a pool of funds to support the expenses for early career researchers. Among universities in the United Kingdom, there is an ongoing commitment to promoting open access publishing by encouraging submission to open access repositories and by assisting researchers in the payment of APCs for immediate open access publication.^13^ At the funder level, more agencies could embrace the Gates Foundation or the Charity Open Access Fund model utilized by the Wellcome Trust, where researchers supported by these funders can request coverage of any associated APCs.^48,49^ Individual researchers can also increasingly choose to release their research open access through venues such as pre-print servers, like *medRxiv*, without undermining their ability to publish their findings in peer-reviewed journals.^50^ Furthermore, so-called “Green Open Access” policies, where researchers can elect to post peer-reviewed papers in open access repositories, are available for many journals, although most researchers do not utilize this option.^51–53^

It is important to note that limiting the amount of science that exists behind a paywall can also have clear advantages for individual researchers and the public.^43,54^ Open access publishing can enhance equity by improving the ability of researchers, either working in low-resource settings or at institutions that cannot support the hefty cost of journal subscriptions, to access publications.^52,55^ Articles published open-access can receive a citation boost compared to those behind paywalls, a boon for researchers looking to increase the audience and impact of their work.^56^ Furthermore, APCs often serve an important purpose in the publication process. APCs can be used to pay the salaries of journal editors, who are often responsible for screening a large number of manuscript submissions, identifying and soliciting appropriate peer-reviewers (and performing their own peer-review), and helping improve the quality of studies as they transition from submission to eventual publication. Moreover, APCs could be used to pay peer reviewers for their efforts - a service currently provided by researchers for free even in cases where researchers are paying thousands of dollars to publish an article.^57^ However, if APCs continue to increase, questions will continue to be raised about journals’ potential profit motives, predatory journals, and hybrid journals that receive payments from both institutions and researchers (we refer to this as ‘double-dipping’). Scientific publishing is changing and it will be necessary for all stakeholders to adapt.

### Limitations of this study

This study is subject to certain limitations. First, we apologize for some of the nomenclature used in our manuscript (i.e., “general medical researchers” and “high-impact medical researchers”). We recognize the limitations of focusing on journal impact factors, and do not believe that authors should be classified as “general” or “high-impact” based upon *one* senior author research or review article published in one of the ten highest impact factor medical journals. Perhaps, we should have classified authors as having “good” or “bad” H-indices or Altmetric Attention Scores, since there seems to be universal agreement about the calculation and use of those metrics.

Second, our estimates do not represent the actual APCs that the index researchers in our sample paid. Without access to the financial records from the index researchers and journals in our sample, we had to make several assumptions about the nature of APC payments, most fundamentally that it was the index author who paid the APCs, rather than a funder or other organization. In particular, articles for which the index researcher was a middle author were excluded, as we assumed index researchers are less likely to pay associated APCs as a middle author. We also did not account for situations in which APCs may have been paid by co-primary or co-senior authors. Additionally, we used the most recent APCs listed on journal website, which may not represent the APCs paid in 2019. For our primary analysis, we assumed that researchers in our sample did not pay the optional APCs for open access publications in hybrid journals. Using publicly available information, it is difficult to determine if open access publications in hybrid journals were paid for by researchers or were available open access due to funder requirements or journal discretion. Furthermore, we did not account for any unlisted discounts or fee waivers provided by journals to researcher institutions in our analyses. Although the true minimum APCs per researcher may be lower than our estimate, our results did not change substantially when analyses were repeated using the lowest APCs listed by journals (excluding waivers). Overall, our sensitivity analyses provide a range of what researchers are likely to have paid.

Third, although Scopus provides a comprehensive accounting of a given researcher’s publication history, not all manuscripts published by a researcher may be indexed on Scopus. Furthermore, Scopus may create multiple researcher profiles for the same researcher, due to changing institutions or different permutations of the researcher’s name. However, we attempted to identify and include all researcher profiles for each index researcher. Second, we relied on articles classified as ‘articles’ or ‘reviews’ on Scopus. Although this method allowed us to objectively screen and classify index researcher articles, it is possible that we may have included or excluded articles that were incorrectly classified by Scopus.

### Life is no picnic

To draw on the picnic analogy mentioned above, the model of open access publication with APCs clearly has great advantages for picnic eaters – who get free food – and for picnic site owners, who can set the charges for food provision. Researchers, however, are left with the effort and cost of growing the food and bringing it to the picnic site. They must then wait for it to be inspected by a team of (usually unpaid) food tasters and be willing to respond to any disparaging comments that these reviewers think fit to make. If they are then allowed on to the picnic site, they are then charged for the privilege of laying out their meal for the public.

Our Twitter analysis provides evidence that this process can take its toll on all but the saintliest contributors. We found that a significant number are driven to exceed the bounds of polite professional discourse. This leads us to postulate that beneath the obvious advantages of open access publication there may lie hidden moral and financial harms to contributors. These could include arguments with colleagues at work, financial arguments with spouses who are unable to appreciate the subtleties of the APC system, sleep disturbance, and even the possible use of alcohol before tweeting.

### Conclusion

This cross-sectional analysis suggests that clinical medical researchers paid as much as $34676 [£27186] in total APCs for their first and senior author research and review articles in 2019. As journals with APCs become more common, it is important to understand the cost to researchers, especially those who may not have the funding or institutional resources to cover these costs. If we do not address rising publication costs, we risk creating a pay-to-publish system that favors well-resourced authors from well-resources institutions and areas of the world. Furthermore, researchers may increasingly turn to Twitter to complain about the financial costs of publishing, and demand compensation for peer-reviewing articles.

## Data Availability

The dataset will be made available via a publicly accessible repository on publication.

## ACKNOWLEDGEMENTS

### Contributors

JDW and JSR first conceived the study idea when arguing about who would have to pay the APC for one of their previous manuscripts. MKE, KN, JSR, and JDW designed this study. MKE, XS, and JJS acquired the author, journal, and APC data. JDW contributed to data collection by spending a full day searching and browsing Twitter (a normal day at the office). MKE conducted the statistical analysis. MKE, JSR, and JDW drafted the manuscript. All authors participated in the interpretation of the data and critically revised the manuscript for important intellectual content, not that there is anything all that important here. MKE and JDW had full access to all the data in the study and take responsibility for the integrity of the data and the accuracy of the data analysis. MKE and JDW are guarantors. JDW provided supervision, and despite being the senior author, begged JSR to pay the APCs.

### Funding

None. Therefore, the APC for this manuscript will come from JDW’s discretionary piggy bank. The authors assume full responsibility for the accuracy and completeness of the ideas presented.

### Competing interests

All authors have completed the ICMJE uniform disclosure form at www.icmje.org/coi_disclosure.pdf and declare: In the past 36 months, XS received a scholarship from China Scholarship Council. KN thinks the estimated APCs are peanuts in comparison to the millions of dollars her employer spends on journal subscriptions, and wonders why authors who avoid APC-induced apoplexy by publishing in subscription journals so infrequently self-archive their papers. RL discloses that he is an avid fan of picnics. JSR is a former Associate Editor of *JAMA Internal Medicine*, a current Research Editor at *BMJ*, pays thousands of dollars annually in APCs, and has received research support through Yale from Johnson and Johnson to develop methods of clinical trial data sharing, from the FDA to establish a Center for Excellence in Regulatory Science and Innovation (CERSI) at Yale University and the Mayo Clinic (U01FD005938), from the Medical Device Innovation Consortium as part of the National Evaluation System for Health Technology (NEST), from the Agency for Healthcare Research and Quality (R01HS022882), from the National Heart, Lung and Blood Institute of the National Institutes of Health (NIH) (R01HS025164, R01HL144644), and from the Laura and John Arnold Foundation. JDW received research support through the Collaboration for Research Integrity and Transparency from the Laura and John Arnold Foundation and through the Center for Excellence in Regulatory Science and Innovation (CERSI) at Yale University and the Mayo Clinic (U01FD005938).

### Patient consent

Not required

### Ethical approval

Not required

### Data sharing

The dataset will be made available via a publicly accessible repository on publication:

### Transparency

The lead author (manuscripts guarantor) (JDW) affirms that this manuscript is an honest, accurate, and transparent account of the study being reported; that no important aspects of the study have been omitted; and that any discrepancies from the study as planned (and, if relevant registered) have been explained.

### License

The Corresponding Author has the right to grant on behalf of all authors and does grant on behalf of all authors, a worldwide license to the Publishers and its licensees in perpetuity, in all forms, formats and median (whether known now or created in the future), to i) publish, reproduce, distribute, display and store the Contribution, ii) translate the Contribution into other languages, create adaptations, reprints, include within collections and create summaries, extracts and/or, abstracts of the Contribution, iii) create any other derivative work(s) based on the Contribution, iv) to exploit all subsidiary rights in the Contribution, v) the inclusion of electronic links from the Contribution to third party material where-ever it may be located; and, vi) license any third party to do any or all of the above. The default license, a CC BY NC license, is needed. This is an Open Access article distributed in accordance with the Creative Commons Attribution Non Commercial (CC BY-NC 4.0) license, which permits others to distribute, remix, adapt, build upon this work non-commercially, and license their derivative works on different terms, provided the original work is properly cited and the use is non-commercial.

See: http://creativecommons.org/licenses/by-nc/4.0/.

## References

1. Ioannidis JPA, Boyack KW, Klavans R. Estimates of the Continuously Publishing Core in the Scientific Workforce. Amaral LANunes, ed. PLoS ONE. 2014;9(7):e101698. doi:10.1371/journal.pone.0101698

2. UNESCO. UNESCO science report: towards 2030. Published online 2015.

3. Rice DB, Raffoul H, Ioannidis JPA, Moher D. Academic criteria for promotion and tenure in biomedical sciences faculties: cross sectional analysis of international sample of universities. BMJ. Published online June 25, 2020:m2081. doi:10.1136/bmj.m2081

4. NSF. Publication Output: U.S. Trends and International Comparisons. Published online 2019.

5. Björk B-C, Solomon D. Open access versus subscription journals: a comparison of scientific impact. BMC Med. 2012;10(1):73. doi:10.1186/1741-7015-10-73

6. Munafò MR, Nosek BA, Bishop DVM, et al. A manifesto for reproducible science. Nat Hum Behav. 2017;1(1):0021. doi:10.1038/s41562-016-0021

7. Resnick B. The war to free science. Vox. Published June 3, 2019. Accessed July 15, 2020. https://www.vox.com/the-highlight/2019/6[cited/3/18271538/open-access-elsevier-california-sci-hub-academic-paywalls

8. Romaine SB Barbara Albee, & Sion. Deal or No Deal | Periodicals Price Survey 2019. Library Journal. Accessed July 23, 2020. https://www.libraryjournal.com?detailStory=Deal-or-No-Deal-Periodicals-Price-Survey-2019

9. Noorden RV. THE TRUE COST OF SCIENCE PUBLISHING. Nature. 2013;495:426–429.

10. Solomon DJ, Björk B-C. Publication fees in open access publishing: Sources of funding and factors influencing choice of journal. J Am Soc Inf Sci Technol. 2012;63(1):98–107. doi:10.1002/asi.21660

11. Directory of Open Access Journals. Directory of Open Access Journals. Accessed February 17, 202 0. https://www.doaj.org/about

12. Dallmeier-Tiessen S, Darby R, Goerner B, et al. Highlights from the SOAP project survey. What Scientists Think about Open Access Publishing. ArXiv11015260 Cs. Published online January 28, 2011. Accessed July 15, 2020. http://arxiv.org/abs/1101.5260

13. Universities UK. Monitoring the Transition to Open Access: December 2017. Universities UK; :52.

14. Nicholas D, Rodríguez-Bravo B, Watkinson A, et al. Early career researchers and their publishing and authorship practices: ECRs publishing and authorship practices. Learn Publ. 2017;30(3):205–217. doi:10.1002/leap.1102

15. Ginther DK, Schaffer WT, Schnell J, et al. Race, Ethnicity, and NIH Research Awards. Science. 2011;333(6045):1015–1019. doi:10.1126/science.1196783

16. Hoppe TA, Litovitz A, Willis KA, et al. Topic choice contributes to the lower rate of NIH awards to African-American/black scientists. Sci Adv. 2019;5(10):eaaw7238. doi:10.1126/sciadv.aaw7238

17. Lawson S. Fee Waivers for Open Access Journals. Publications. 2015;3(3):155–167. doi:10.3390/publications3030155

18. Peterson A, Emmett A, Greenberg M. Open Access and the Author-Pays Problem: Assuring Access for Readers and Authors in the Global Academic Community. J Librariansh Sch Commun. 2013;1(3):eP1064. doi:10.7710/2162-3309.1064

19. InCites Journal Citation Reports. Accessed June 16, 2020. https://jcr.clarivate.com/JCRLandingPageAction.action?Init=Yes&SrcApp=IC2LS&SID=H2-BzzoBH7fIIbTAafOrld9xxFix2F5DwRnQm6-18x2dSruBm8K4GbN6ZPQtUZbfUAx3Dx3DADCSreBC9mJb3O7KgOE12wx3Dx3D-qBgNuLRjcgZrPm66fhjx2Fmwx3Dx3D-h9tQNJ9Nv4eh45yLvkdX3gx3Dx3D

20. World Health Organization. WHO Regional Offices. Accessed April 20, 2020. https://www.who.int/about/who-we-are/regional-offices

21. Elsevier. Elsevier/Impactstory agreement will make open access articles easier to find on Scopus. Elsevier Connect. Accessed July 15, 2020. https://www.elsevier.com/connect/elsevier-impactstory-agreement-will-make-open-access-articles-easier-to-find-on-scopus

22. Laakso M, Björk B-C. Hybrid open access—A longitudinal study. J Informetr. 2016;10(4):919–932. doi:10.1016/j.joi.2016.08.002

23. Article Processing Charges | Wolters-Kluwer. Accessed July 20, 2020. https://wkauthorservices.editage.com/open-access/hybrid.html

24. Article Processing Charges | Oxford. Oxford Academic. Accessed July 20, 2020. https://academic.oup.com/journals/pages/ita-journals-a-to-z

25. Article Processing Charges | Karger. Accessed July 20, 2020. https://www.karger.com/Journal/Index?oa=true&sub=true&dis=false&s=acta%20cytologica&ss=&max=5&skip=0&open=null&closeSF=null

26. Article Processing Charges | Taylor & Francis Online. Accessed July 20, 2020. https://www.tandfonline.com/openaccess/openjournals

27. Article Processing Charges | SAGE Publications Ltd. Accessed July 20, 2020. https://uk.sagepub.com/en-gb/eur/pure-gold-open-access-journals-at-sage

28. Article Processing Charges | Future Medicine. Accessed July 20, 2020. https://www.futuremedicine.com/authorguide/openaccess

29. Article Processing Charges | Hindawi. Hindawi. Accessed July 20, 2020. https://www.hindawi.com/publish-research/authors/article-processing-charges/

30. Article Processing Charges | MAG Online Library. Accessed July 20, 2020. https://www.magonlinelibrary.com/page/authors/openaccess

31. Article Processing Charges | MDPI. Accessed July 20, 2020. https://www.mdpi.com/apc#Journal_Specific_APCs

32. Article Processing Charges | Thieme Open. Accessed July 20, 2020. https://open.thieme.com/web/19/apcs

33. Article Processing Charges | Emerald Publishing. Accessed July 20, 2020. https://www.emeraldgrouppublishing.com/services/authors/publish-us/publish-open-access/journal#apc-charges

34. Article Processing Charges | JMIR. Accessed July 20, 2020. https://www.jmir.org

35. Article Processing Charges | Springer. www.springer.com. Accessed July 20, 2020. https://www.springer.com/gp/open-access/springer-open-choice

36. Article Processing Charges | Elsevier. Accessed July 20, 2020. https://www.elsevier.com/about/policies/pricing

37. Article Processing Charges | SAGE Open. SAGE Publications Ltd. Published October 6, 2015. Accessed July 20, 2020. https://uk.sagepub.com/en-gb/eur/sage-choice-journal-and-pricing-exceptions

38. Falagas ME, Zarkali A, Karageorgopoulos DE, Bardakas V, Mavros MN. The Impact of Article Length on the Number of Future Citations: A Bibliometric Analysis of General Medicine Journals. Fortunato S, ed. PLoS ONE. 2013;8(2):e49476. doi:10.1371/journal.pone.0049476

39. Yearly Average Currency Exchange Rates | Internal Revenue Service. Accessed July 23, 2020. https://www.irs.gov/individuals/international-taxpayers/yearly-average-currency-exchange-rates

40. Shamseer L, Moher D, Maduekwe O, et al. Potential predatory and legitimate biomedical journals: can you tell the difference? A cross-sectional comparison. BMC Med. 2017;15(1):28. doi:10.1186/s12916-017-0785-9

41. Björk B-C, Solomon D. Article processing charges in OA journals: relationship between price and quality. Scientometrics. 2015;103(2):373–385. doi:10.1007/s11192-015-1556-z

42. Shaw DM, Elger BS. Unethical Aspects of Open Access. Account Res. 2018;25(7-8):409-416. doi:10.1080/08989621.2018.1537789

43. Laakso M, Björk B-C. Anatomy of open access publishing: a study of longitudinal development and internal structure. BMC Med. 2012;10(1):124. doi:10.1186/1741-7015-10-124

44. Khoo SY-S. Article Processing Charge Hyperinflation and Price Insensitivity: An Open Access Sequel to the Serials Crisis. Liber Q. 2019;29(1):1. doi:10.18352/lq.10280

45. Pollock D, Michael A. Open access mythbusting: Testing two prevailing assumptions about the effects of open access adoption. Learn Publ. 2019;32(1):7–12. doi:10.1002/leap.1209

46. Shieber SM. Equity for Open-Access Journal Publishing. PLoS Biol. 2009;7(8):e1000165. doi:10.1371/journal.pbio.1000165

47. “Plan S” and “cOAlition S” – Accelerating the transition to full and immediate Open Access to scientific publications. Accessed July 15, 2020. https://www.coalition-s.org/

48. Baptista D. Charity Open Access Fund (COAF) open access spend and compliance monitoring: 2017-18. Published online 2019:2664082 Bytes. doi:10.6084/M9.FIGSHARE.8010617

49. Open Access Policy. Published January 1, 1AD. Accessed July 15, 2020. https://www.gatesfoundation.org/How-We-Work/General-Information/Open-Access-Policy

50. Massey DS, Opare MA, Wallach JD, Ross JS, Krumholz HM. Assessment of Preprint Policies of Top-Ranked Clinical Journals. JAMA Netw Open. 2020;3(7):e2011127–e2011127. doi:10.1001/jamanetworkopen.2020.11127

51. Baffy G, Burns MM, Hoffmann B, et al. Scientific Authors in a Changing World of Scholarly Communication: What Does the Future Hold? Am J Med. 2020;133(1):26–31. doi:10.1016/j.amjmed.2019.07.028

52. Smith E, Haustein S, Mongeon P, Shu F, Ridde V, Larivière V. Knowledge sharing in global health research – the impact, uptake and cost of open access to scholarly literature. Health Res Policy Syst. 2017;15. doi:10.1186/s12961-017-0235-3

53. Button OA. Open Access Button. Accessed July 23, 2020. https://openaccessbutton.org

54. Cuschieri S. WASP: Is open access publishing the way forward? A review of the different ways in which research papers can be published. Early Hum Dev. 2018;121:54–57. doi:10.1016/j.earlhumdev.2018.02.017

55. Chan L, Kirsop B, Arunachalam S. Towards Open and Equitable Access to Research and Knowledge for Development. PLOS Med. 2011;8(3):e1001016. doi:10.1371/journal.pmed.1001016

56. The Open Access Citation Advantage Service (OACA). SPARC Europe. Accessed July 15, 2020. https://sparceurope.org/what-we-do/open-access/sparc-europe-open-access-resources/open-access-citation-advantage-service-oaca/

57. Diamandis EP. Peer review as a business transaction. Nature. 2015;517(7533):145–145. doi:10.1038/517145a

